# A Cross-Sectional Survey on Smartphone Usage Pattern, the Level of Mobile Phone Dependence and Psychosocial Effects among Undergraduate Students in a Malaysian University

**DOI:** 10.1101/2020.01.06.20016592

**Authors:** Nisha Syed Nasser, Loh Jia Ling, Aida Abdul Rashid, Hamed Sharifat, Umar Ahmad, Buhari Ibrahim, Salasiah Mustafa, Hoo Fan Kee, Ching Siew Mooi, Subapriya Suppiah

**Affiliations:** Centre for Diagnostic Nuclear Imaging, Faculty of Medicine and Health Sciences, Universiti Putra Malaysia, 43400 Serdang, Selangor, Malaysia; Department of Imaging, Faculty of Medicine and Health Sciences, Universiti Putra Malaysia, 43400 Serdang, Selangor, Malaysia; Medical Genetics Laboratory (MGL), Genetics and Regenerative Medicine Research Centre, Faculty of Medicine and Health Sciences, Universiti Putra Malaysia, 43400 Serdang, Selangor, Malaysia; Department of Anatomy, Faculty of Basic Medical Sciences, Bauchi State University, PMB 65, Gadau, Nigeria; Department of Physiology, Faculty of Basic Health Sciences, Bauchi State University, PMB 65, Gadau, Nigeria; Neurology Unit, Department of Medicine, Faculty of Medicine and Health Sciences, Universiti Putra Malaysia, 43400 Serdang, Selangor, Malaysia; Department of Family Medicine, Faculty of Medicine and Health Sciences, Universiti Putra Malaysia, 43400 Serdang, Selangor, Malaysia

**Keywords:** Smartphone Addiction Scale, education, social networking, Malaysia

## Abstract

**Objective:** Problematic smartphone use (PSU) is the development of pathological dependence at the expense of performing activities of daily living, thus having a negative health and psychosocial impact on the users. Previous PSU studies focused on medical students and little is known regarding its effect on students undergoing other fields of study. The objective of this study is to identify the pattern of smartphone usage and determine the psychosocial factors affecting PSU among undergraduate students in Malaysia and compare the pattern among different fields of study.

**Method:** A prospective cross-sectional study was conducted using validated Smartphone Addiction Scale–Malay version (SAS-M) questionnaire. One-way ANOVA was used to determine the correlation between the patterns of smartphone usage among the students categorised by their ethnic groups, hand dominance and by their field of study. MLR analysis was applied to predict PSU based on socio-demographic data, smartphone usage patterns, psychosocial factors and field of study.

**Results:** A total of 1060 students completed the questionnaire. The majority of students had PSU (60.7%). Students used smartphones predominantly to access SNAs, namely Instagram. Longer duration on the smartphone per day (≥ 9 hours), age at first using a smartphone and depression carried higher risk of developing PSU, whereas the field of study (science vs. arts based) did not contribute to an increased risk of developing PSU.

**Conclusion:** Findings from this study can help better inform university administrators about at-risk groups of undergraduate students who may benefit from targeted intervention designed to reduce their addictive behavior patterns.

## INTRODUCTION

Smartphone usage among university students, albeit beneficial in many aspects, has lead to problematic overdependance in this digital era. It has increased exponentially over the past 10 years.^1^ It is estimated that close to 2.32 billion people in the worldwide population own a smartphone, whilst in Malaysia, the latest statistics in 2014 recorded 10.31 million active smartphone users i.e. close to 89.4% of the Malaysian population use their smartphones to access the Internet.^2, 3^ Problematic smartphone use (PSU) can be defined as an overdependence on smartphones and the inability to regulate its use, despite experiencing ill-effects and unwelcome consequences in daily living.^4, 5^ The features of PSU are closely related to the criteria for Internet Gambling Disorder as stated in the Diagnostic and Statistical Manual of Mental Disorders, 5th edition (DSM-5). Despite this, the addicted individuals persist with their behaviour; ignoring adverse consequences and display escalation of the behavior and demonstrate withdrawal symptoms when they attempt to reduce their addictive behaviour.^6, 7^Numerous somatic and psychosocial ill-effects have been associated with PSU. Specifically, it has been associated with musculoskeletal disorders affecting the fingers/hands/wrist due to the excessive use of the devices.^8^ In fact, smartphone overuse is known to lead to sleep problems, and somatic effects such as fatigue, tension, headache and dizziness and social problems.^9^ Furthermore, a study conducted in South Korea reported that the social problems caused by PSU were similar to the features of other behavioural addictions.^10^

Several tools have been developed to measure the severity of PSU, in particular questionnaires have been developed to measure Internet addiction by Kimberly Young in 1998, and a revised version in the Chinese language developed by Chen et al in 2003, which have been adapted to assess for smartphone addiction or problematic smartphone use.^11, 12^ More recently, a group of Korean researchers have modified these questionnaires to suit mobile devices overuse and to assess for PSU.^13^ This was the Smartphone Addiction Scale (SAS) questionnaire that was later translated into the Malay language and validated by Ching et al. (2015) to suit the Malaysian population, called the Smartphone Addiction Scale-Malay version (SAS-M) questionnaire.^14^ Other tools that have been utilised to assess for Internet and smartphone addiction or problematic use include functional magnetic resonance imaging (fMRI) studies that look into the brain regions affected by smartphone overuse.^15^

It is important to note that Malaysia represents a multiethnic country in South East Asia, however, due to cultural adaptations and adjustments, the various ethnic groups share many similarities based on Asian values and can be considered homogeneous in their smartphone usage behaviours. Furthermore, when focusing on undergraduate university students, it is known that they share almost similar education background and IQ levels. Nevertheless, much of previous studies on Internet and smartphone usage among young adults have focused on medical students.^16, 17, 18^

There is limited data available regarding PSU among students from different fields of study other than the medical field.^19^ So far there has been no similar data available in a Malaysian population. Therefore, we conducted a survey among undergraduate students from a Malaysian public university to determine the pattern of smartphone use among undergraduate students from various fields, and the psychosocial effects associated with PSU. We also investigated the students’ perception as to how their smartphone usage affected their academic performance. We were particularly interested to correlated the pattern of usage with the degree of PSU; using the SAS-M questionnaire as a tool to gauge the severity of dependence.

## MATERIALS AND METHOD

### Subjects Recruitment

After acquiring ethical clearance from the local institutional committee (JKEUPM) (UPM/TNCPI/RMC/1.4.18.2), a prospective cross-sectional study was conducted among the undergraduate students in Universiti Putra Malaysia (UPM) using convenience sampling method. The respondents were recruited via advertisements in student notice boards and also through communication with undergraduate course lecturers. The sample size calculation was based on a prevalence study conducted among the student population.^20^ The sample size formula is:

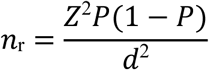

with the prevalence of smartphone addiction, =0.084 (16), =1.96 and (precision of an estimate) of 0.0168 generated a required sample size of 1048. In anticipation of potential dropouts in the study, we distributed 1362 copies of the questionnaires. Participation in this survey was voluntary and the students gave informed consent to participate in the study, in keeping with the Declaration of Helsinki principles. No incentive was provided to the respondents and the students were made clear that their responses or denial to join this study would not affect their grades.

### Tool for data collection

#### Smartphone Addiction Scale-Malay version questionnaire (SAS-M)

We employed the SAS-M questionnaire to determine the severity of PSU among the undergraduate students. SAS-M is a validated 33-point questionnaire which was rated using a 6-point Likert scale, ranging from 1 to 6 (1=strongly disagree to 6=strongly agree). This well-structured, self-administered questionnaire was designed and validated in the Malay language by Ching et al. in 2015. The ROC curve gave an optimal AUC of 0.801 (95% CI = 0.746 to 0.855) and best cut-off score for identifying at-risk cases was ≥ 98, giving a sensitivity of 71.43%, specificity of 71.03%, PPV of 64.10% and NPV of 77.44%.^14^ Based upon the cut-off score suggested, students with SAS-M scores of ≥ 98 were considered to have PSU and those with scores < 98 were considered to be using their smartphone at a non-hazardous level.^14^

#### Depression, Anxiety and Stress scale (DASS-21)

In addition to the SAS-M questionnaire, the subjects also completed a validated 21-item self-reported DASS-21 questionnaire, which was used to evaluate the state of emotional response i.e. depression, anxiety, and stress.^21^ Each of these was rated on a four-point Likert scale. The scores ranged from 0=did not apply to them at all to 4=applied to them most of the time. The following cut-off scores were used for each subscale: depression: normal 0–9, mild 10–13, moderate 14–20, severe 21–27 and extremely severe 28+; anxiety: normal 0–7, mild 8–9, moderate 10–14, severe 15–19 and extremely severe 20+; stress: normal 0–14, mild15–18, moderate 19–25, severe 26–33 and extremely severe 34+.

### Data Analyses

#### Statistical Analysis

The data was analyzed using SPSS V25.0 (SPSS 25.0, Chicago, IL, USA). General demographic data of the students was described using tables for categorical data, and medians and ranges for continuous variables. Comparison of continuous variables was performed using the Independent-samples t test. Pearson’s correlation was done to determine the strength of the relationship between SAS-M scores and pattern of smartphone usage (years of smartphone use and duration of smartphone use per day). A p value < 0.05 was considered statistically significant. One-way ANOVA was used to determine the correlation between the patterns of smartphone usage among the students categorised by their ethnic groups, hand dominance and by their field of study. Multilinear regression analysis (MLR) was applied to predict PSU based on socio-demographic data, smartphone usage patterns, psychosocial factors and field of study.

## RESULTS

We collected back 1060 completed sets with a response rate of approximately 78%. The respondents, ranging from 17 – 25 years old, from various faculties in UPM completed and returned the questionnaires, having a female preponderance ratio of 3:1 as shown in Table I. A major portion of the students were from the Faculty of Medicine and Health Sciences (FMHS) (35.8%); followed by 10.8% of students from the Faculty of Veterinary Medicine, 13.0% from the Faculty of Engineering, 9.8% from the Faculty of Agriculture, 9.0% from the Faculty of Modern Languages and Communication, and the rest of the faculties had smaller percentage of participants. The respondents were grouped into 3 main fields of study i.e. medicine (n=380, 35.8%), applied sciences (n=423, 39.9%), and social sciences (n=257, 24.2%) as shown in Table I.

**Table I:**
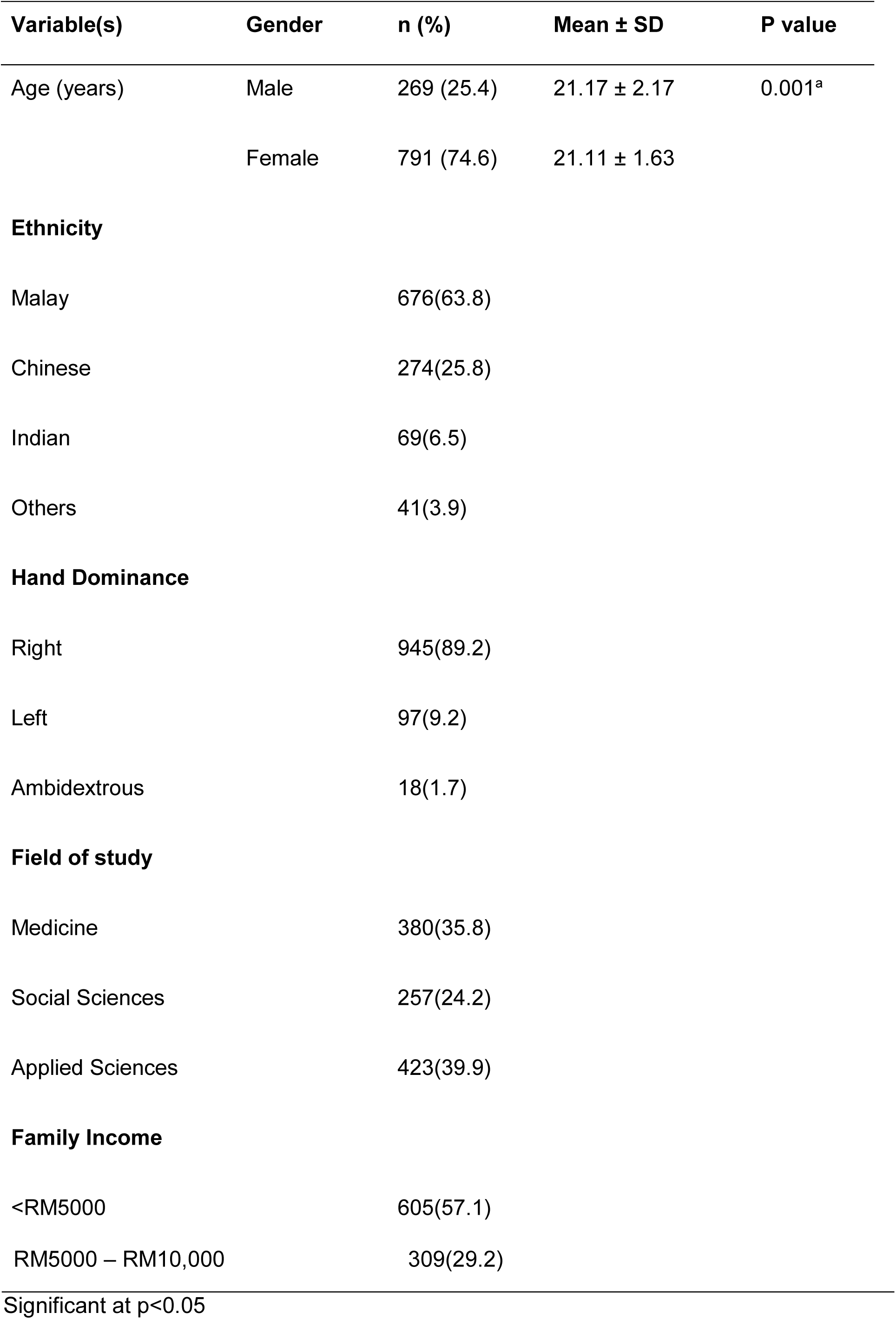
Socio-demographic characteristics of respondents (n = 1060)

### Pattern of smartphone usage

The mean age of students when they first started using smartphones was 13.7 years old; male students (14.20 ± 2.67) and female students (13.52 ± 2.63). Additionally, the number of years using a smartphone was significantly greater among the female students (6.65 ± 2.90) as compared to male students (5.80 ± 2.79), p-value<0.001. On average, the respondents spent 8 hours per day using their smartphones. Additionally, female students stayed on their smartphones significantly longer (8.47 hours, SD=5.19) as compared to the male students (6.94 hours, SD=4.53), p-value<0.001. The Malay students used their smartphones for a longer time (8.87 ± 5.16) compared to Chinese (6.31 ± 4.32), Indian (7.16 ± 4.71) and other races (8.52 ± 5.66) (p<0.05).. One-way ANOVA did not reveal any significant differences in the age of first using smartphones, duration of using a smartphone and the average time spent per day using a smartphone, in terms of hand dominance. Additionally, using one-way ANOVA, significant differences were noted among Medicine, Social Sciences and Applied Sciences students for age of first using smartphones and duration of using a smartphone (p<0.05). Average time spent per day using smartphone did not differ significantly among the three field of study groups as demonstrated in Table II.

**Table II:** Pattern of smartphone usage among respondents (n=1060) stratified by field of study.

| | Faculty | Min-Max | Mean ± SD | F statistic | P value* |
| --- | --- | --- | --- | --- | --- |
| Age of first using | Medicine | 6 - 21 | $14.06 \pm 2.84$ | 6.820 | 0.001* |
| smartphone (years) | Social Sciences | 76 - 19 | $13.29 \pm 2.47$ | | |
|  | Applied Sciences | 76 - 21 | 13.61 ± 2.55 |  |  |
| Years of using a smartphone | Medicine | 1 - 18 | 6.29 ± 2.94 | 4.536 | 0.011* |
|  | Social Sciences | 1 - 16 | 6.12 ± 2.93 |  |  |
|  | Applied Sciences | 2 - 19 | 6.75 ± 2.82 |  |  |
| Average time spent per day in smartphone (hours) | Medicine | 1 - 24 | 7.63 ± 4.92 | 2.456 | 0.086 |
|  | Social Sciences | 1 - 24 | 8.25 ± 5.41 |  |  |
|  | Applied Sciences | 1 - 20 | 8.39 ± 4.96 |  |  |

\*Significant at $p < 0.05$
One-way ANOVA

### Purpose of smartphone usage

The topmost reason for using smartphones across all fields of study was for social networking and communications (66.8%) followed by 21% for studies related purposes as illustrated in Figure 1a. Overall, the most widely used smartphone application was noted to be WhatsApp (72.0%) followed by Instagram (13.7%) and Facebook (7.5%) as shown in Figure 1b. The majority of students from all faculties (>70%) used WhatsApp as the preferred smartphone application, which is mainly utilised for communication as the posts are not publicly available to users outside of the WhatsApp chat group. Hence, Instagram was noted to be the most popular social networking application (SNA) among the students and has exceeded Facebook usage by approximately 6%.

**Fig. 1:**
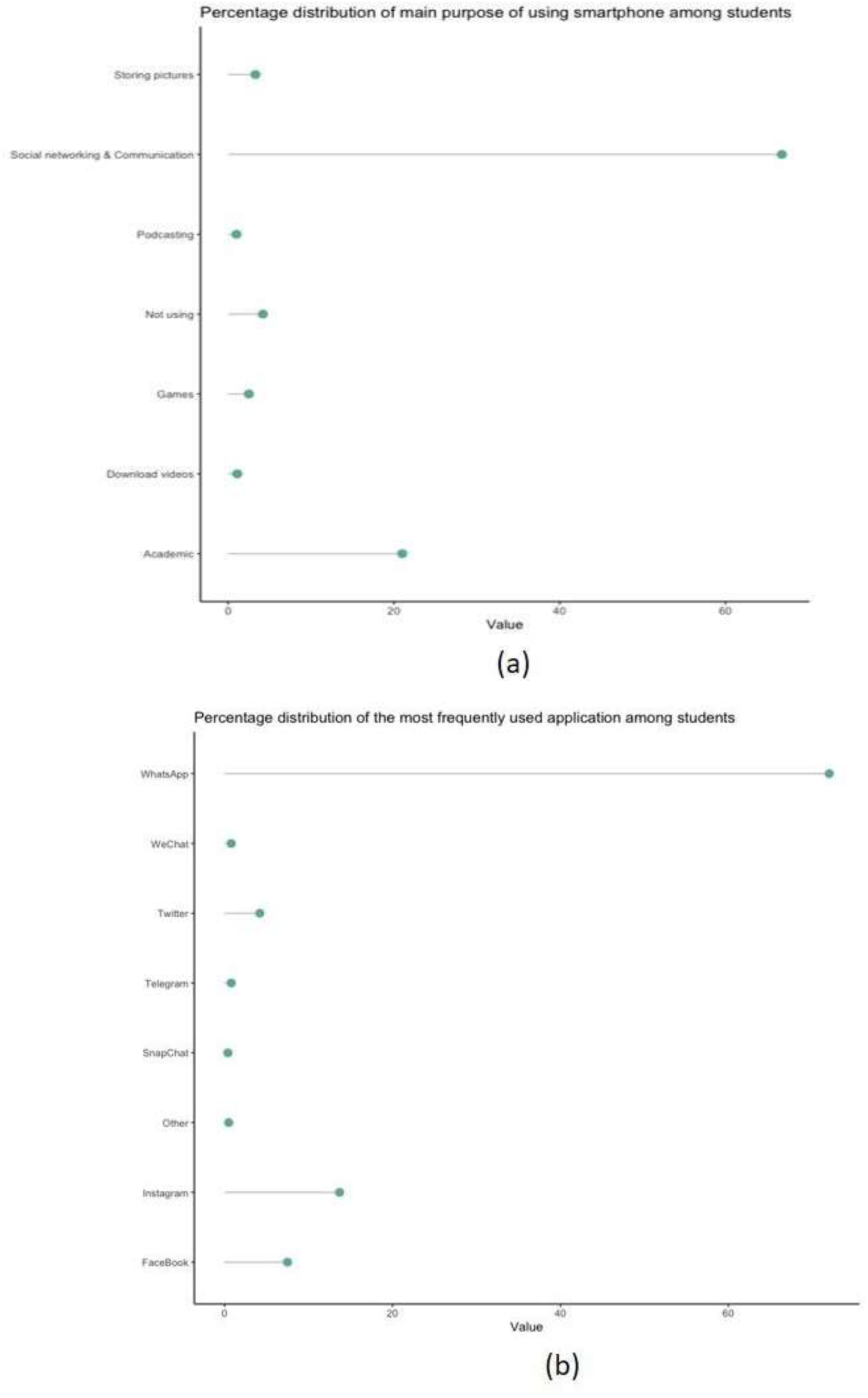
Pattern of smartphone usage among undergraduate students. (a) The distribution of main purpose of using smartphones. (b) The most frequently used application on smartphones.

### Prevalence of problematic smartphone use and factors affecting it

The prevalence of PSU among the undergraduate students in this public Malaysian university was 60.7% comprising of 643 students (males n=165; female n= 478) out of 1060 total respondents. The remaining 417 students were identified to have healthy level of smartphone usage (males n=104; females n=313). Among the PSU students identified in this study, 212 were Medical students (33%), 159 were Social Sciences students (24.7%) and 272 were Applied Sciences students (42.3%). Overall, we noted a higher prevalence of PSU among students within the Applied Sciences field (64.3%), compared to 61.9% in Social Sciences and 55.8% of students within the Medicine group as shown in Figure 2.

**Fig. 2:**
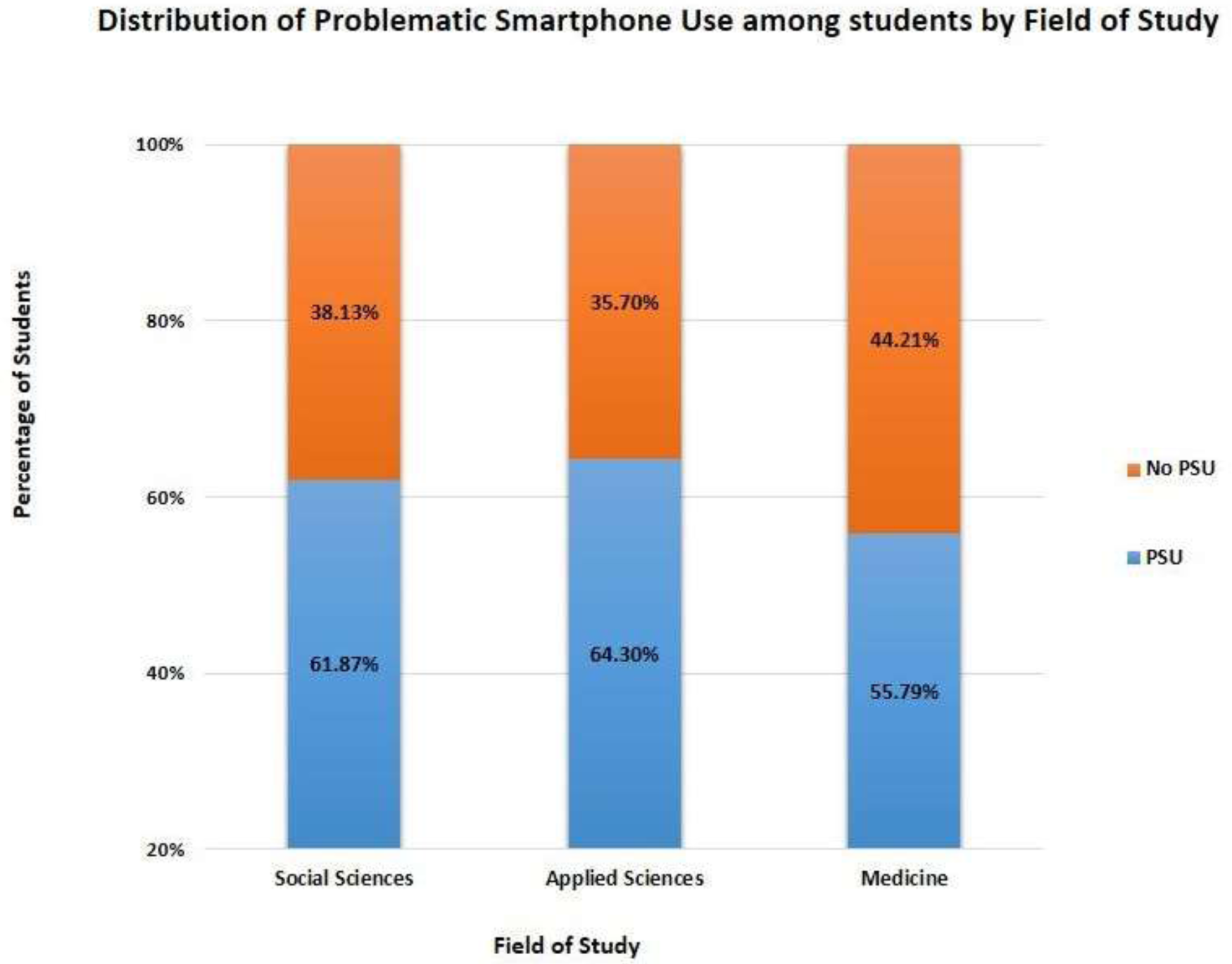
Distribution of problematic smartphone use among students based on the field of study.

### Effects of smartphone usage on the perception of academic performance

Almost half (49.62%) of the students reported that the use of smartphone did not have any effect on their studies, whilst 30.28% reported that smartphones had assisted in improving their academic performance. The remaining 20.09% of students reported that smartphones had caused deterioration in their study performance. Students who perceived that they had deterioration in their academic performance had significantly higher SAS-M scores (110.39 ± 27.63) as opposed to students who stated that they had experienced improvement in their studies performance (102.89 ± 25.16). Overall, 30.3% of students declared that they experienced improvement in their academic performance that is attributed to their smartphone usage, as opposed to 49.6% who declared it did not affect their performance and 20.1% who deemed that they had deterioration in their academic performance.

### Predicting smartphone addiction from socio-demographic factors, smartphone usage pattern, and psychological factors

Most of our subjects did not suffer from any significant depressive symptoms (53%) and none had major depression. Nevertheless, approximately 17% and 20% of the respondents reported that they had mild and moderate depression respectively. About 5% of them reported that they had experienced severe depressive symptoms. Furthermore, approximately 8%, 31% and 13% of the respondents reported that they had mild, moderate and severe anxiety. Notably, 15% of our respondents declared mild stress symptoms and 9% were found to have moderate stress levels.

To test the hypothesis that socio-demographic factors, smartphone usage pattern, field of study and psychological factors can account for a significant proportion of variance in SAS-M score, multiple linear regression (MLR) was performed. Four models regression analysis were used to predict factors that influenced PSU. Total scores for depression was entered in Model 1; The average time spent per day using smartphone was entered in Model 2; Age of using smartphone was entered in Model 3 and total scores for anxiety was entered in Model 4.

From Table III, age of first using smartphone, the average time spent per day in smartphone, anxiety, and depression were the only variables that can be included in the multivariable analysis. The four model summary table from the output results were obtained and model 4 was chosen because 31.4% of the model variation is explained by the independent variables compared to the remaining models 1, 2, 3 with 24.2%, 30.1% and 30.8% respectively. Note that the larger the R and R^2^ the more accurate we predict SAS-M.

**Table III:** MLR analysis for predicting SAS-M score.

| Variable | Unadjusted<br>(Simple regression) |  | Adjusted<br>(Multiple regression) |  |  |
| --- | --- | --- | --- | --- | --- |
| | B ($\beta$, 95%CI) | p value | B ($\beta$, 95%CI) | p value | VIF |
| Age | -0.384<br>(-1.419, 0.650) | 0.466 | - | - | - |
| Age of first using<br>smartphone (years) | -0.858<br>(-1.537, -0.179) | 0.013 | -0.944<br>(-1.590, -0.298) | 0.004 | 1.321 |
| Years of using a<br>smartphone | -0.389<br>(-1.036, 0.258) | 0.238 | -0.485<br>(-1.075, -0.106) | 0.107 | 1.313 |
| Average time spent<br>per day in<br>smartphone (hours) | 0.887<br>(0.590, 1.184) | <0.001 | 0.887<br>(0.590, 1.183) | <0.001 | 1.013 |
| Stress | 0.177<br>(-0.230, 0.464) | 0.509 | - | - | - |
| Anxiety | 0.265<br>(-0.054, 0.585) | 0.104 | 0.320<br>(0.033, 0.607) | 0.029 | 2.225 |
| Depression | 0.504<br>(0.189, 0.820) | 0.002 | 0.554<br>(0.280, 0.827) | <0.001 | 2.210 |
| Sciences-based<br>studies | 3.287<br>(-0.451, 7.205) | 0.085 | 2.928<br>(-0.657, 6.513) | 0.109 | 1.0611 |
| F value |  |  | 20.073 |  |  |
| P-value |  |  | 0.001 |  |  |
| Adj R<sup>2</sup> |  |  | 0.098 |  |  |

Based on the prediction model 4 from MLR, the formula to predict SAS-M estimation is SAS-M = 100.943 + [-0.944 ′ age of first using smartphone (AFS)] _+_ [0.887 ′ average time spent per day in smartphone (ATS)] + [0.320 ′ Anxiety] + [0.554 ′ Depression]

## DISCUSSION

To the best of our knowledge, this is the first study conducted in Malaysian involving a large population of young adults from a diverse field of study. Furthermore, the changes in cultural practice among Asian students who have undergone urbanization is apparent, as we detected a high prevalence of PSU i.e. 60.7% among the undergraduate university students. This is comparable to findings among adolescents in other more developed countries such as England (10%), Switzerland (16.9%), Germany (23.4%), Saudi Arabia (71.9%) and South Korea (25.5%).^22-26,10^ Pertaining to PSU, the mean SAS-M scores of our PSU students was 103.3 (±26.02), and was noted to be lower than the South Korean population who had a mean score of 110.02 but much higher compared to college students in Turkey who had a mean score of 75.68.^13, 27^

Our respondents used smartphones for an average of 8 hours per day, which is comparable to studies in other countries like the United States that noted average usage time of 8-10 hours daily ^28^. This finding agreed with the observation made by Haug et al. in 2015, which noted that longer duration of smartphone use in a typical day predisposed young adults in Switzerland to have PSU.^23^ We noted a higher likelihood of developing PSU when students used their devices for 9 hours and more per day, as opposed to non-addicted students who used their devices for a mean duration of 7 hours (p<0.001).

Due to prolonged duration of use, a whopping 48.3% of our respondents reported that they had poor sleep quality and experienced daytime tiredness due to their late-night smartphone overuse. This was almost similar to an observation done among Lebanese students, whereby 35-38% of them experienced similar symptoms that were detrimental to their health.^29^ Thus, we recommend that students limit their daily smartphone usage to 7 hours or less to prevent addictive behaviours. Methods that can be utilized include switching off the mobile data and wireless fidelity (WiFi) connection when spending social time with friends and family, during driving and when going to bed.

The majority (69.7%) of our students reported that they used smartphones mainly for social networking purposes followed by for communication purposes. WhatsApp, which was the most commonly utilised mobile application, has gained popularity as an effective communications tool due to its ease to send message, provides group sharing of information and is cost-effective in enabling the users to share images and videos with each other. Nevertheless, the commonest purpose to be online among these students was noted to be for accessing SNA. This may be due to the availability and ease of various SNAs like Facebook, Instagram and Snapchat, which provide an outlet for the students’ socializing needs. The most widely used SNAwas Instagram (13.6%) and Facebook (7.5%). This shows the evolution of pattern usage since 2016 whereby another Malaysian study had reported that there was a high Facebook addiction rate of 47% among their students.^30^ Instagram users were double the amount of Facebook users in our study population. We postulate that this could be due to the youth who are more inclined to use Instagram because they find it more fun in the aspects of photo-sharing, and sharing their ‘daily moments’ with their friends and family who follow them; without having to worry about other people ‘stealing’ their pictures or videos, as they cannot be downloaded and saved directly without the owner’s permission. In fact, one of our students responded that *‘Facebook is for the older generation’* and *‘Most of the younger people now use Instagram’*.

There was a significant difference in SAS-M scores among students who perceived that they experienced deterioration in their studies compared to those who improved in their studies, which coroborated with results from previous studies. However, this is a new observation compared to the results reported by Boumosleh & Jaalok (2017) which had stated that there was no significant correlation between academic performance and PSU.^29^ In this instance, students may benefit from using their level of academic performance and its deterioration, as a cue to warn them regarding their addictive behaviours pertaining to smartphone use. Students may try to seek advice from counselors or academic advisors to overcome this problem. Study by Long et al. (2016) among Chinese undergraduate students identified students majoring in Humanities to have increased risk of developing PSU.^19^ In contrast our study did not identify science based study to be a significant predictor of developing PSU. Thus, developing risk of PSU is age related rather than the field of study.

The main limitation of this study is the dependency on self-reported data. If we were able to access more primary information, it could provide a more accurate picture of the actual pattern of usage among the students. Furthermore, regarding usage of social networking services, more information is needed with regards to the nature of their interactions, e.g. the type of photos uploaded on Instagram and whether other types of addiction such as whether substance abuse mediated this. There is also a potential to delve further into the effects of peer pressure and conformation to social norms among the youth of this millennium in future studies.

## CONCLUSION

Proper utilization of smartphones must be inculcated among the students to prevent any untoward effects and academic deterioration. Findings from this study can help better inform university administrators about at-risk groups of undergraduate students who may benefit from targeted intervention designed to reduce their addictive behavior patterns.

## Data Availability

The data that support the findings of this study are available from the corresponding author upon request.

## ACKNOWLEDGEMENTS

This study was supported by the research grant GP-9549800 and GP-IPS/9580800. The authors would like to thank the students and lecturers from different faculties of the university who helped to disseminate and respond to the questionnaires.

